# Real-world study of the effectiveness of BBIBP-CorV (Sinopharm) COVID-19 vaccine in the Kingdom of Morocco

**DOI:** 10.1101/2022.04.23.22274112

**Authors:** Yaowen Zhang, Jihane Belayachi, Yunkai Yang, Qiang Fu, Lance Rodewald, Hongling Li, Bing Yan, Ying Wang, Yanna Shen, Qian Yang, Weiyun Mu, Rong Tang, Chen Su, Tianfang Xu, Majdouline Obtel, Abdelkader Mhayi, Rachid Razine, Redouane Abouqal, Yuntao Zhang, Xiaoming Yang

## Abstract

The Kingdom of Morocco approved BBIBP-CorV (Sinopharm) COVID-19 vaccine for emergency use on 22 January 2021 in a two-dose, three-to-four-week interval schedule. We conducted a case-control study to determine real-world BBIBP-CorV vaccine effectiveness (VE) against serious or critical hospitalization of individuals RT-PCR-positive for SARS-CoV-2 during the first five months of BBIBP-CorV use in Morocco.

The study was conducted among adults 18-99 years old who were tested by RT-PCR for SARS-CoV-2 infection between 1 February and 30 June 2021. RT-PCR results were individually linked with outcomes from the COVID-19 severe or critical hospitalization dataset and with vaccination histories from the national vaccination registration system. Individuals with partial vaccination (<2 weeks after dose two) or in receipt of any other COVID-19 vaccine were excluded. Unadjusted and adjusted VE estimates against hospitalization for serious or critical illness were made by comparing two-dose vaccinated and unvaccinated individuals in logistic regression models, calculated as (1-odds ratio) * 100%.

There were 348,190 individuals able to be matched across the three databases. Among these, 140,892 were fully vaccinated, 206,149 were unvaccinated, and 1,149 received homologous BBIBP-CorV booster doses. Unadjusted, full-series, unboosted BBIBP-CorV VE against hospitalization for serious or critical illness was 90.2% (95%CI: 87.8% - 92.0%). Full-series, unboosted VE, adjusted for age, sex, and calendar day of RT-PCR test, was 88.5% (95%CI: 85.8% - 90.7%). Calendar day- and sex-adjusted VE ranged from 93.9% to 100% for individuals <60 years, and was 53.3% for individuals 60 years and older. There were no serious or critical illnesses among BBIBP-CorV-boosted individuals.

Effectiveness of Sinopharm’s BBIBP-CorV was consistent with phase III clinical trial results. Two doses of BBIBP-CorV was highly protective against COVID-19-associated serious or critical hospitalization in working-age adults under real-world conditions and moderately effective in older adults. Booster dose VE should be evaluated, as booster doses of BBIBP-CorV are recommended and are being used.

## BACKGROUND

COVID-19 is a public health emergency of international concern. As of March 2022, the World Health Organization (WHO) has received reports of more than 475 million cases and 6 million COVID-19 deaths ^[1]^. The first case of COVID-19 in the Kingdom Morocco was reported to WHO on 2 March 2020. As of 8 February 2022, there have been 1,147,243 COVID-19 cases and 15,593 COVID-19 associated deaths reported by Morocco to WHO ^[1]^. There have been two complete epidemic waves in Morocco: the first started early November 2020, and the second started in August 2021. In January 2022, case counts rose again, signaling onset of a third wave.

In April 2021, more than 90% of SARS-CoV-2 isolates were Alpha variants, with the rest being Beta and Gamma. In August 2021, during the second wave, 80% were Delta and 20% were Alpha. In January 2022, during the current third wave, 90% of isolates have been Omicron, with the remainder being Delta ^[2]^. The scientific consensus is that while COVID-19 vaccines prevent some SARS-CoV-2 infections, they are most effective at prevention of COVID-19 related hospitalization, serious illness, and death.

COVID-19 vaccines were first introduced to Morocco on 28 January 2021 and recommended for individuals 18 years and above. Thus far, there have been five COVID-19 vaccines in widespread use in Morocco: inactivated virus vaccine (Beijing CNBG’s BBIBP-CorV), virus vectored vaccines (AstraZeneca’s Vaxzevria and Gamaleya’s Gam-COVID-Vac), mRNA vaccine (Pfizer-BioNTech’s Comirnaty), and modified virus vector vaccine (Janssen’s Ad26.COV 2-S). As of 12 January 2022, there have been 51,321,365 doses of COVID-19 vaccines administered to 24,701,243 people; 66.14% of the total population has had one or more doses, and 61.83% of the population (23,088,979 people) has been fully vaccinated ^[3]^.

BBIBP-CorV is a whole-virus, inactivated, alum-adjuvanted vaccine; its safety and immunogenicity were demonstrated in pre-clinical and phase I and II clinical trials^[4-6]^. Efficacy of BBIBP-CorV for preventing symptomatic infection was 78.1% in a phase III study conducted in United Arab Emirates, Bahrain, Egypt, and Jordan^[7]^. WHO approved BBIBP-CorV for emergency use listing on 5 May 2021^[8]^; Morocco approved BBIBP-CorV for emergency use on 22 January 2021^[9]^, and was one of the first countries to approve the vaccine. According to the Morocco Department of Health, citizens and foreigners residing in Morocco could be vaccinated with BBIBP-CorV vaccine upon appointment. Priority groups for vaccination were healthcare personnel, front-line workers, seniors, and patients with chronic diseases. Full primary vaccination with BBIBP-CorV requires two intramuscular injections separated by an inter-dose interval of 21 to 28 days^[10]^.

The phase III clinical trials of BBIBP-CorV had too few severe/critical cases to provide reliable efficacy estimates against the most severe outcomes^[7]^. Real-world studies with larger sample sizes are needed for such assessments. The protective effectiveness of BBIBP-CorV against serious or critical hospitalization associated with COVID-19 had not been assessed previously in Morocco. We report a real-world study to estimate the effectiveness of BBIBP-CorV in the Kingdom of Morocco using official surveillance, testing, and vaccination data during the vaccine’s first five months of use in Morocco.

## METHODS

### Study design

We conducted a case-control study to evaluate vaccine effectiveness (VE) of BBIBP-CorV against serious or critical COVID-19-associated hospitalization among Morocco residents aged 18-99 who were tested by Reverse Transcription Polymerase Chain Reaction (RT-PCR) for SARS-CoV-2 infection between 1 February and 30 June 2021.

### Data Sources

Study data were obtained from three databases, with individual-level data linked by national identification number. Vaccination data, including vaccination date, and vaccine type and dose were obtained from the National Vaccination Registry (NVR), which is an electronic health record system that records complete vaccination histories. RT-PCR SARS-CoV-2 test results data were obtained from E-labs, which is a Morocco national laboratory network for COVID-19 diagnostic specimens tested by RT-PCR. Clinical data on severity of COVID-19-associated hospitalization were recorded in a public health surveillance dataset made available for this study.

### Outcomes and vaccination status

COVID-19 associated hospitalization for serious or critical illness was defined any hospitalization for severe/critical illness of an individual with SARS-CoV-2 RT-PCR positivity, regardless of whether hospitalization was a direct consequence of COVID-19 infection.

Individuals were classified as fully vaccinated if at least 14 days had passed since the administration of a second dose of BBIBP-CorV vaccine. An unvaccinated, control population consisted of individuals who had not received any doses of any COVID-19 vaccine. Individuals who received three doses of BBIBP-CorV were categorized as boosted.

### Statistical analysis

Means and standard deviations (SD) were used to describe continuous variables; categorical variables were expressed as frequencies and percentages. We estimated VE against serious or critical hospitalization by using logistic regression models. Unadjusted VE and adjusted VE (adjusting for age, sex, and calendar day of RT-PCR test) were calculated as (1-odds ratio) * 100%. All data analyses were performed with SAS software (version 9.4, SAS Institute Inc., Cary, NC, USA).

### Ethical review

The study was approved by the Rabat Ethical Review Committee for Biomedical Research at Mohammed V University. A waiver of informed consent was granted for this observational study. Authorization from the National Commission for the Protection of Personal Data (CNDP) was also obtained.

## RESULTS

Figure 1 shows the study flow diagram and inclusions and exclusions of potential subjects. From 1 February 2021 to 30 June 2021, a total of 436,438 records from the Covid-19 related severe or critical hospitalization dataset and E-labs were able to be matched with vaccination histories in the NVR. After exclusions for missing data on sex or age, administration of a vaccine other than BBIBP-CorV, and partial vaccination, 348,190 records remained and comprised the analytic dataset. There were 140,892 individuals fully vaccinated with two doses of BBIBP-CorV, 206,149 individuals completely unvaccinated against COVID-19, and 1,149 booster-vaccinated individuals (Table 1).

**Figure 1.**
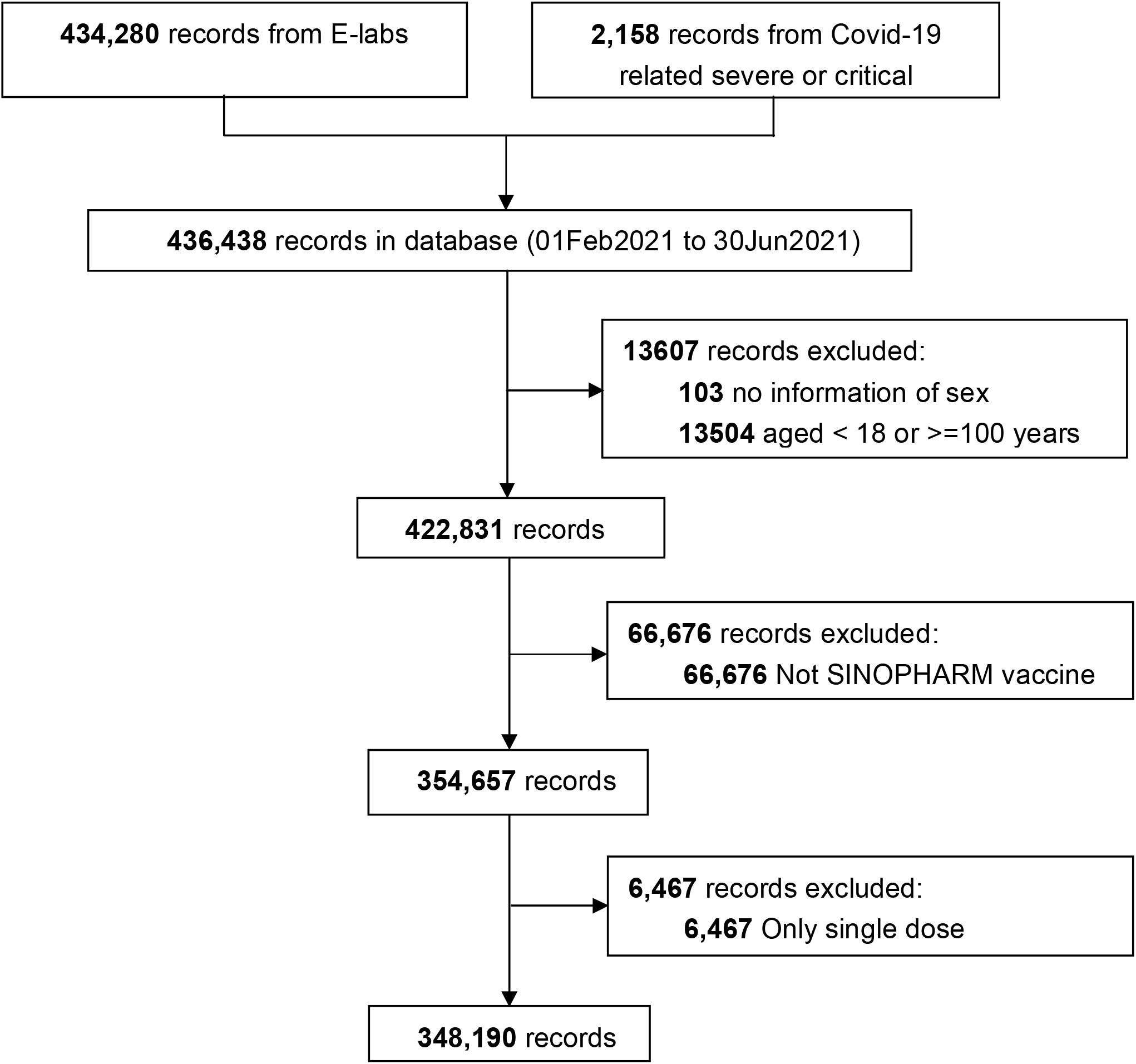
Flowchart of the study.

**Table 1.** Characteristics of the study subjects by BBIBP-CorV vaccination status, Morocco, 1 February 2021 through 30 June 2021.

|  | Not Vaccinated<br>**N=206,149** | Fully Vaccinated<br>**N=140,892** | Booster-vaccinated<br>**N=1,149** | Total<br>**N=348,190** |
| --- | --- | --- | --- | --- |
| Sex, n (%) |  |  |  |  |
| Male | 116,816 (56.7) | 81,580 (57.9) | 1,024 (89.1) | 199,420 (57.3) |
| Female | 89,333 (43.3) | 59,312 (42.1) | 125 (10.9) | 148,770 (42.7) |
| Age (Years) |  |  |  |  |
| Mean (SD) | 40.7 (15.1) | 37.9 (13.5) | 45.1 (15.8) | 39.6 (14.6) |
| Age group (Years), n (%) |  |  |  |  |
| 18-29 | 56,553 (27.4) | 47,313 (33.6) | 201 (17.5) | 104,067 (29.9) |
| 30-39 | 53,558 (26.0) | 35,226 (25.0) | 336 (29.2) | 89,120 (25.6) |
| 40-49 | 41,896 (20.3) | 29,515 (20.9) | 195 (17.0) | 71,606 (20.5) |
| 50-59 | 27,576 (13.4) | 21,263 (15.1) | 151 (13.1) | 48,990 (14.1) |
| ≥60 | 26,566 (12.9) | 7,575 (5.4) | 266 (23.2) | 34,407 (9.9) |

Table 2 shows serious or critical hospitalization broken down by BBIBP-CorV vaccination status, sex, and age group. Compared with unvaccinated individuals, fully and booster-vaccinated individuals had a lower rates of serious or critical hospitalization.

**Table 2.** Serious or critical hospitalization by vaccination status, sex, and age group.

| Age group: | Serious/critical<br>hospitalization<br>N (%) | No serious/critical<br>hospitalization<br>N (%) | p-value |
| --- | --- | --- | --- |
| **Main analysis** |  |  |  |
| Not Vaccinated | 1,345 (0.65) | 204,804 (99.35) | <.001 |
| Fully Vaccinated | 91 (0.06) | 140,801 (99.94) |  |
| Booster-vaccinated | 0 (0) | 1,149 (100) |  |
| **Subgroup analysis** |  |  |  |
| **Male** |  |  |  |
| Not Vaccinated | 688 (0.59) | 116,128 (99.41) | <.001 |
| Fully Vaccinated | 60 (0.07) | 81,520 (99.93) |  |
| Booster-vaccinated | 0 (0) | 1,024 (100) |  |
| **Female** |  |  |  |
| Not Vaccinated | 657 (0.74) | 88,676 (99.26) | <.001 |
| Fully Vaccinated | 31 (0.05) | 59,281 (99.95) |  |
| Booster-vaccinated | 0 (0) | 125 (100) |  |
| **18-29 years** |  |  |  |
| Not Vaccinated | 65 (0.11) | 56,488 (99.89) | <.001 |
| Fully Vaccinated | 0 (0) | 47,313 (100) |  |
| Booster-vaccinated | 0 (0) | 201 (100) |  |
| **30-39 years** |  |  |  |
| Not Vaccinated | 182 (0.34) | 53,376 (99.66) | <.001 |
| Fully Vaccinated | 0 (0) | 35,226 (100) |  |
| Booster-vaccinated | 0 (0) | 336 (100) |  |
| **40-49 years** |  |  |  |
| Not Vaccinated | 254 (0.61) | 41,642 (99.39) | <.001 |
| Fully Vaccinated | 6 (0.02) | 29,509 (99.98) |  |
| Booster-vaccinated | 0 (0) | 195 (100) |  |
| **50-59 years** |  |  |  |
| Not Vaccinated | 353 (1.28) | 27,223 (98.70) | <.001 |
| Fully Vaccinated | 18 (0.08) | 21,245 (99.90) |  |
| Booster-vaccinated | 0 (0) | 151 (100) |  |
| **>=60 years** |  |  |  |
| Not Vaccinated | 491 (1.85) | 26,075 (98.15) | <.001 |
| Fully Vaccinated | 67 (0.88) | 7,508 (99.12) |  |
| Booster-vaccinated | 0 (0) | 266 (100) |  |

Unadjusted, unboosted full-series BBIBP-CorV vaccine effectiveness against serious or critical hospitalization was 90.2% (95%CI: 87.8% - 92.0%). After adjusting for age, sex, and calendar day of RT-PCR test in the logistic regression model, adjusted full-series, unboosted VE against serious or critical hospitalization was 88.5% (95%CI: 85.8% - 90.7%). Adjusted full-series, unboosted VE was higher among females than males (92.6% vs 83.8%) and among 18-59-year-olds than individuals 60 years and older (93.9% to 100% vs 53.3%). There were no serious or critical hospitalizations among boosted individuals (Table 3).

**Table 3.** Unadjusted and adjusted two-dose, unboosted BBIBP-CorV vaccine effectiveness against serious or critical hospitalization among 18-99-year-olds in Morocco.

|  | Unadjusted VE<br>% (95%CI) * | Adjusted VE<br>% (95CI) |
| --- | --- | --- |
| **Main analysis** |  |  |
| Not Vaccinated | Ref. | Ref. |
| Fully Vaccinated | 90.2 (87.8-92.0) | 88.5 (85.8-90.7) |
| **Subgroup analysis** |  |  |
| **Male** |  |  |
| Not Vaccinated | Ref. | Ref. |
| Fully Vaccinated | 87.6 (83.8-90.5) | 83.8 (78.9-87.6) |
| **Female** |  |  |
| Not Vaccinated | Ref. | Ref. |
| Fully Vaccinated | 92.9 (89.9-95.1) | 92.6 (89.4-94.9) |
| **18-29 years** |  |  |
| Not Vaccinated | Ref. | Ref. |
| Fully Vaccinated | 100 (NA-NA) | 100 (NA-NA) |
| **30-39 years** |  |  |
| Not Vaccinated | Ref. | Ref. |
| Fully Vaccinated | 100 (NA-NA) | 100 (NA-NA) |
| **40-49 years** |  |  |
| Not Vaccinated | Ref. | Ref. |
| Fully Vaccinated | 96.3 (92.5-98.5) | 97.0 (93.3-98.7) |
| **50-59 years** |  |  |
| Not Vaccinated | Ref. | Ref. |
| Fully Vaccinated | 93.5 (89.5-95.9) | 93.9 (90.2-96.2) |
| **>=60 years** |  |  |
| Not Vaccinated | Ref. | Ref. |
| Fully Vaccinated | 52.6 (38.7-63.3) | 53.3 (39.6-63.9) |

Abbreviations: VE, vaccine effectiveness; CI, confidence interval; NA, value is not available.
\*Unadjusted VE and adjusted VE (adjusting for age, sex, and calendar day of RT-PCR test) were calculated as (1-odds ratio) \* 100% from the logistic regression model.

## DISCUSSION

This nationwide, real-world study in Morocco showed that unadjusted effectiveness of two-dose, primary BBIBP-CorV vaccination against serious or critical COVID-19 associated hospitalization was 90.2%. When adjusted for age, sex, and calendar date, two-dose VE against serious or critical hospitalization was 88.5%. Adjusted protective effectiveness of two doses of BBIBP-CorV against serious or critical hospitalization among 18-60-year-olds ranged from 93.9% to 100% and was 53.3% among adults 60 and older. No serious or critical hospitalizations occurred among booster dose recipients, but there were too few booster dose recipients for logistic regression analysis.

Our VE estimates are consistent with efficacy estimates from overseas phase III clinical trials of BBIBP-CorV ^[7]^ and overseas observational VE studies. In Argentina, two-dose vaccine effectiveness (VE) against COVID-19 related mortality was 84% among those aged ≥60 years ^[11]^. In a large cohort of health care workers in Peru, two-dose VE was 50% against infection and 94% against COVID-19-related mortality ^[12]^. In a nation-wide observational study in Hungary with 895,000 adult BIBP-CorV recipients, two-dose VE was 69% against infection and 88% against COVID-19 related mortality ^[13]^. In a cohort study conducted in the United Arab Emirates among individuals 15 years and older, two-dose BBIBP-CorV VE was 79.8% against hospitalization ^[14]^.

Our study results are consistent with other real world studies of inactivated COVID-19 vaccines. In Chile, full-series CoronaVac VE was 87.5% against hospitalization and 90.3% against ICU admission ^[15]^. In Brazil, during a P.1 variant epidemic, full-series CoronaVac was 59.0% effective against hospitalizations and 71.4% effective against death ^[16]^. Also in Brazil, in a setting with high SARS-CoV-2 Gamma variant transmission, CoronaVac effectiveness against symptomatic infection of health care workers was 56.2% ^[17]^.

Pfizer-BioNTech’s BNT162b2 mRNA vaccine and Moderna’s mRNA-1273 vaccine have been well studied around the world. Full-series vaccination with BNT162b2 or mRNA-1273 VE against severe COVID-19 is 90% or more in the general population. Two studies in Israel showed BNT162b2 VE against severe disease to be 92% and 97.5% ^[18,19]^. In the United States, BNT162b2 or mRNA-1273 VE against ICU admission was 90%^[20]^. In Canada, BNT162b2 VE was 98% against hospital admission or death^[21]^. Our results showed that BBIBP-CorV had similar VE against severe disease in the overall population.

A cohort study in the UAE^[14]^ with 176,640 individuals aged ≥ 15 years showed that two-dose BBIBP-CorV VE was 92.2% for preventing COVID-19-related critical care admission when compared to a non-vaccinated group – a study design and finding similar to ours. Wu^[22]^ and Kang^[23]^ performed real-world studies on the effectiveness of inactivated vaccines used in China against illness caused by the Delta variant in outbreaks in Henan and Guangdong provinces, respectively. Wu and colleagues found that among adults ≥ 18 years, two-dose inactivated VE against severe or critical COVID-19 was 92% ^[22]^, and Kang and colleagues found that two-dose inactivated VE was 100% ^[23]^ against severe illness. The two studies were too small to estimate VE separately for BBIBP-CorV and CoronaVac, the two inactivated vaccines in use in the outbreak settings.

Our study in Morocco found that VE against serious or critical hospitalization was lower among older adults compared with working age adults - VE ranged from 94% to 100% among younger adults but was 53% among those 60 years and over. Effectiveness Pfizer, Moderna, and AstraZeneca COVID-19 vaccines for prevention of severe disease or hospitalization varied from 70% to 90% among older adults ^[21, 24-29]^. A test negative case-control study conducted in Brazil^[30]^ showed that the effectiveness of CoronaVac vaccine against hospital admission in older adults was 55.5%. A study in the Netherlands^[31]^ showed that Spikevax vaccine (mRNA-1273) effectiveness against ICU admissions declined with age and was 34% among individuals ≥70 years of age. Moustsen-Helms^[32]^ and Yelin^[33]^ showed results that are similar to ours. There are several possible reasons for decreased effectiveness with older age. Elderly people have more comorbidities^[33]^, and vaccine-induced immune responses in the elderly are slower and SARS-CoV-2 neutralization is lower^[34]^. Vaccination of elderly individuals is critically important. All COVID-19 vaccines, including BBIBP-CorV, now require a booster dose to maintain protection over time and across age groups. Protective VE must be carefully monitored during real-world use of COVID-19 vaccines.

Our study has several limitations. First, there were no analyses of virus strains, therefore BBIBP-CorV VE against specific SARS-CoV-2 variants could not be estimated. Second, we did not have individual-level comorbidity data, precluding subgroup analysis of VE. Third, we did not have data on reasons for critical hospitalization. As a result, our study could not distinguish critical hospitalization caused by SARS-CoV-2 infection from critical hospitalization coincidental to SARS-CoV-2 infection. This limitation may partially explain the lower VE among older subjects, who are more likely to be hospitalized than younger subjects. For example, Emborg^[35]^ and colleagues found that BNT162b2 mRNA VEs against all-cause hospital admissions and COVID-19-related hospital admissions among ≥65-year-olds were 37% and 87%, respectively. Fourth, the timing of the study was such that too few individuals had received booster doses, precluding an analysis of BBIBP-CorV booster dose VE.

In conclusion, consistent with the results of phase III clinical trials, BBIBP-CorV vaccine is highly protective against severe and critical hospitalization in real-world conditions. VE was found to be greater in working age adults than elderly adults. Monitoring VE is important for adjusting vaccine recommendations.

## Data Availability

All data produced in the present study are available upon reasonable request to the authors.

